# Science and knowledge translation strategies to the public during health emergencies: systematic review of RCTs

**DOI:** 10.1101/2025.07.21.25331954

**Authors:** Melody Taba, Michael Anthony Fajardo, Eliza Ferguson, Rachael Keast, Jocelyne M Basseal, Kirsten McCaffery, Carissa Bonner

## Abstract

**Introduction:** Effective science and knowledge translation is essential during public health emergencies. During the COVID-19 pandemic, rapidly evolving research had to be translated to the public under challenging conditions.

**Objectives:** This review aimed to identify randomised trials of COVID-19 science and knowledge translation strategies targeting the public and evaluated their effectiveness in improving psychological, behavioural and/or health outcomes.

**Methods:** A literature search was done across PubMed, Embase, Scopus, CINAHL, and PsycINFO in July 2023 and November 2024. Studies were screened and extracted according to PRISMA guidelines. Interventions reporting behavioural outcomes were coded using the Behaviour Change Technique (BCT) taxonomy and the Cochrane risk-of-bias tool was used to assess study quality.

**Results:** Of 345 records screened, 48 eligible studies were included. Most were online experiments testing message framing, with a smaller number conducted in applied settings such as health professional–delivered education. Significant positive effects were reported in most studies; 30 out of 40 studies with psychological outcomes (e.g. knowledge), 28 out of 40 studies with behavioural outcomes (e.g. intention to mask). Only one study measured a health outcome, with no significant effect. Effective features commonly included video and animation formats and messages from health experts and credible sources. The most frequent BCTs were ‘information about health consequences’ (33 studies) and ‘credible source’ (19 studies). Risk of bias was low in 42 studies.

**Conclusions:** These findings highlight a diverse range of strategies that improved outcomes during the COVID-19 pandemic. Better use of behavioural science taxonomies and core outcome sets could help researchers advance the field further during future emergencies.

**PROSPERO registration number:** CRD42023446093.

**ARTICLE SUMMARY:** *Strengths and limitations of this study:*

- This study followed rigorous, pre-registered protocol based on PRISMA guidelines, and included a theory-based behavioural science taxonomy to compare studies in a standardised way.
- Due to the heterogeneity of the studies, meta-analyses were inappropriate for this data.
- The complex interventions and study designs made it difficult to isolate the effects of specific intervention components across studies.
- Inconsistencies in author use of science and knowledge translation terms may have affected the comprehensiveness of the search.
- Most studies measured behavioural intentions rather than actual behaviour, and actual behaviour outcomes were self-reported.

## INTRODUCTION

Communicating scientific evidence to the public is an essential component of public health emergency management. Science and knowledge translation plays a crucial role in policy decisions, behaviour change and maintaining public trust.^1^ This is especially important during emergencies, when uncertainty is high and the need for public action is urgent.

During the early emergency phase of the COVID-19 pandemic (2020-2022), research and understanding of the virus evolved quickly,^2^ creating challenges for effective science translation to the public. The World Health Organization (WHO) also recognised an “infodemic” in early 2020, where the large volume of misinformation online made it difficult to convey reliable scientific information about COVID-19 to the public.^3, 4^ Competing messages, shifting recommendations, and politicisation of science complicated the public’s ability to make evidence-based decisions.^5^

Reflections on the COVID-19 emergency response have underscored the importance of open, transparent communication about evolving scientific evidence, as essential for enabling effective prevention and management.^6, 7^ To inform future emergency responses that depend on large-scale behaviour change, there is an urgent need to evaluate how science translation strategies shaped behavioural outcomes.^3, 8^ A wide variety of COVID-19 translation strategies were employed globally, but there is a need to synthesise what we know about effectiveness across multicomponent interventions. Behavioural science taxonomies can be used to systematically identify the “active ingredients” of complex interventions.^9^

### Objective

The aim of this systematic review was to identify randomised trials of COVID-19 science and knowledge translation strategies targeting the public, and evaluate their effectiveness in improving psychological, behavioural and health outcomes.

## METHODS

This systematic review was proceeded by a rapid scoping review prepared by our research team for the World Health Organization, which identified previous reviews of science and knowledge translation during public health emergencies.^10^ This identified some general categories of strategies including messaging content and community partnerships; however newer technology and media approaches relevant to the COVID-19 response were not well explored at the review level at this time.

A more targeted review focused on randomised studies during COVID-19 was therefore developed, using the guidelines provided by the Preferred Reporting Items for Systematic Reviews and Meta-analysis (PRISMA).^11^ The question was defined as: “In the general public/communities (population), do science translation strategies about COVID-19 information (intervention) improve psychological, behavioural and health outcomes (outcomes) compared to usual care (control) in randomised controlled studies (study design)?” The protocol was registered on PROSPERO (CRD42023446093).

### Inclusion and Exclusion Criteria

Studies were considered eligible if they met the following criteria:

1. Published about or during the emergency phase of the COVID-19 pandemic, from 2020 onwards, in peer reviewed journals.
2. *Population:* Targeted at the general public or specific public communities
3. *Intervention:* Science and knowledge translation content or processes to explain science relating to COVID-19, including virus spread and prevention measures.
4. *Comparators:* Usual processes or care.
5. *Outcomes:* Report quantitative psychological, behavioural, or health outcomes.
6. *Study Design:* Randomised controlled studies (including variations of randomisation e.g., cluster, stepped-wedge, crossover).

Studies where the target intervention group were health professionals or policy makers were excluded, as the objective of this research was to focus on science translation for consumers and communities. Only papers published in peer-reviewed journals were included. Studies that did not gather empirical data (i.e., review, meta-analysis) were not excluded via search parameters as the intention was to identify and evaluate original research articles. If a review was identified, its included papers were cross-referenced with the final inclusion list to identify additional papers. Finally, studies were excluded if they were not available in English language.

### Search Strategy

Database searches of PubMed, Embase, Scopus, CINAHL, and PsycINFO were originally conducted in July 2023 and updated in November 2024. The search terms were based on refinements from our earlier scoping review, with input from science communication and infectious disease experts (See Online supplemental file 1). Additionally, reference lists from studies that met inclusion criteria for the current systematic review were examined, as well as previous reviews of knowledge translation interventions for COVID-19 in the community.

### Data management

Five researchers were involved in data review (MT, MF, EF, RK, CB). Search results were imported into Covidence for screening. Duplicates were removed, and two researchers screened titles/abstracts separately (MT, MF). Studies that did not meet criteria were excluded. Two reviewers screened full texts (MT, MF) and assessed them against eligibility criteria. Data extraction was completed in Microsoft Excel by two researchers (MT, MF) (See Online supplemental file 2). Discrepancies between reviewers were resolved through consensus, or discussion with a third author (CB). For the meta-analysis, 12 corresponding authors were contacted for additional data, and 4 authors provided data. Meta-analysis was conducted in SPSS 29.

#### Quantitative data

We extracted study design, country the study took place, COVID-19 context (i.e., public health and social measures taken, COVID-19 health impacts in study country), sample characteristics (age, gender, race/ethnicity), sample size, comparison groups (usual processes/care as usual), psychological measures (e.g., knowledge of, attitudes towards, and beliefs about COVID-19 vaccines), behavioural measures (e.g., engagement in preventative behaviours, willingness to be vaccinated), health outcomes (e.g., self-reported infections), and measures of effect (e.g., odds ratio, relative risk, difference in means/proportions) comparing intervention and comparator groups with a 95% confidence interval.

#### Intervention content data

We extracted the study’s definition of knowledge translation and intervention format/source/content/framing. A behaviour change technique (BCT) taxonomy^9^ was applied to studies with behavioural outcomes. Two psychologists (RK, EF) completed web-based training to apply the taxonomy to published methods and used a predefined data extraction form to extract data. Data was checked by a third author trained in psychology and behaviour change theory (CB), and discrepancies were resolved through discussion.

#### Risk of bias (quality) assessment

We used Version 2 of the Cochrane risk-of-bias tool for randomized trials (RoB-2).^12^ Studies were critically appraised independently by two review authors (MT, MF) using the relevant Cochrane RoB-2 tools for the study design. Disagreements were resolved by discussion.

#### Patient and public involvement

This research was done without patient or public involvement.

## RESULTS

### 1. Summary of included studies

The search identified 345 articles related to community-based COVID-19 science and knowledge translation strategies (Figure 1). Study characteristics are in Table 1, summaries in Table 2 (further study details in Online supplemental file 6). The studies were conducted across over 19 countries; the most frequently represented were the USA (n=17), China (n=5) and UK (n=3), and six studies collected data from multiple countries.^13–18^ Most studies focussed on general adult populations, some aiming for nationally representative samples (e.g., USA: Agley et al., 2023; Bokemper et al., 2022, Thunstorm et al., 2021;^19–21^ UK: Egan et. al, 2021, Freeman et al., 2021;^22, 23^ Australia: Bonner et. al, 2024; Li, 2024^24, 25^), whilst others focused on specific eligibility criteria like unvaccinated individuals,^26–33^ vaccine-hesitant individuals,^31, 34, 35^ older adults,^33, 36, 37^ parents^13, 31^ or students.^38, 39^ The mean number of years from year of completion of study to publication was 1.63 (SD=0.95) years (6 papers had missing information about year of completion of study).

**Figure 1.**
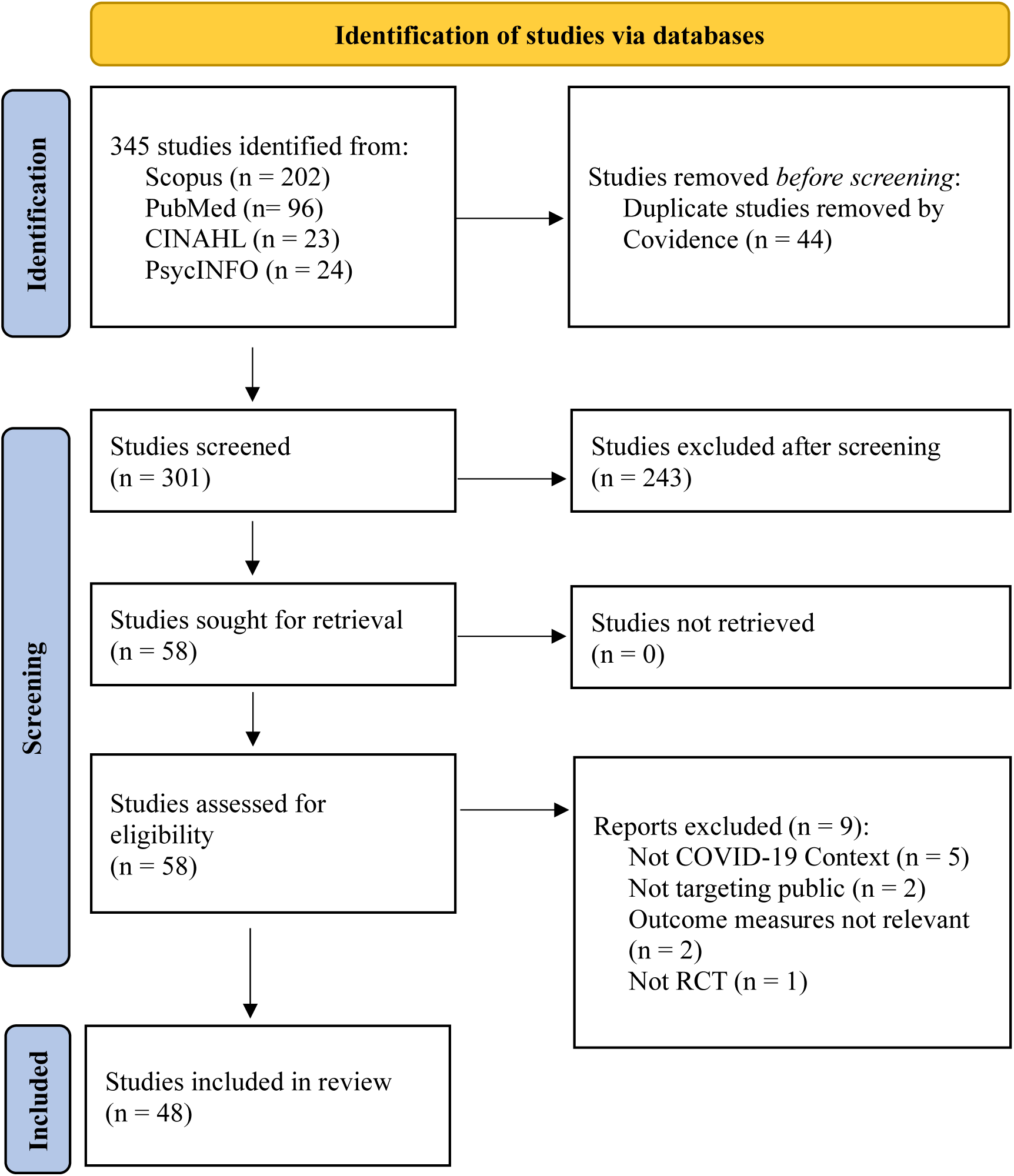
PRISMA (Preferred Reporting Items for Systematic Reviews and Meta-Analyses) diagram.

**Table 1.** Study characteristics (n = 48)

| **Characteristic** | **n** |
| --- | --- |
| **Region** |  |
| Americas (North & South) | 17 |
| Africa | 2 |
| Asia | 13 |
| Europe | 7 |
| Oceania | 3 |
| Global/multiple | 6 |
| **Setting** |  |
| Applied | 15 |
| Online | 33 |
| **Intervention format** |  |
| Text | 28 |
| Video/animation | 11 |
| Infographic | 6 |
| Online (incl. social media) | 5 |
| In person | 3 |
| **Outcome category** |  |
| Psychological | 40 |
| Behavioural | 40 |
| Health | 1 |
| **Psychological subcategory** |  |
| Knowledge | 15 |
| Disease knowledge | 6 |
| Vaccine knowledge | 6 |
| Mask knowledge | 1 |
| Guideline knowledge | 1 |
| Test knowledge | 1 |
| Attitudes and beliefs | 31 |
| Towards vaccine | 14 |
| Towards preventative behaviours | 11 |
| Towards disease | 10 |
| Towards self | 5 |
| Towards misinformation | 5 |
| Towards science | 3 |
| Emotions | 5 |
| Negative emotions | 5 |
| Positive emotions | 1 |
| **Behavioural subcategory** |  |
| Behavioural intentions | 34 |
| Actual behaviour | 9 |
| Vaccination-related behaviour | 19 |
| Other preventative health behaviours | 19 |
| Health information sharing/seeking behaviour | 12 |

**Table 2.** Summary of included studies.

| Study | Sample | Intervention | Effect of intervention on main outcomes |
| --- | --- | --- | --- |
| Abdel-Qader (2022a)<br>*Jordan* | 295 COVID-19 preventative behaviour and vaccine hesitant adults | Virtual coaching by pharmacist via Zoom (vs. no coaching) | Improved COVID-19 preventative behaviours |
| Abdel-Qader (2022b)<br>*Jordan* | 305 COVID-19 vaccine hesitant adults | Virtual coaching by pharmacist and physician via Facebook Live (vs. no coaching) | Improved vaccine hesitancy, knowledge and attitudes towards vaccine, and vaccine rates |
| Abroms (2024)<br>*USA* | 478 unvaccinated adults | Professional-led Facebook group (vs. usual care) | Improved vaccine intentions, responsibility. No change on vaccine rates or confidence. |
| Agley (2023)<br>*USA* | 1,526 adults (nationally representative) | Social media post with cognitive and normative claims (vs. only cognitive claims) | No effect on trust in science |
| Barnawi (2023)<br>*Saudi Arabia* | 508 adults | Educational video (vs. no video) | Improved knowledge and attitudes towards vaccine |
| Beleites (2024)<br>*USA* | 792 adults | Animated story-based video (vs. unrelated video) | Improved knowledge and attitudes towards vaccine, no effect on vaccine or information sharing intentions |
| Bian (2024)<br>*China* | 202 college students and their grandparents | Online health education and SMS reminders (vs. no education) | Improved attitudes, vaccine intentions and rates in grandparents |
| Bokemper (2022)<br>*USA* | 9,263 adults (nationally representative) | Persuasive written messages (Prosocial vs. self-interest; collective vs. individual; recklessness vs. bravery; vs. unrelated message) | Improved attitudes and preventative behaviour intentions (recklessness & collective framing) |
| Bonner (2024)<br>*Australia* | 2,005 adults (nationally representative) | Animated videos about risk as probability or chance or graph (vs. usual care) | Improved knowledge of COVID-19 and vaccinations (videos) |
| Buttenheim (2024)<br>*South Africa* | 7,118 unvaccinated adults | Persuasive written messages (social norm vs. convenience, freedom vs. solution to pandemic) (vs. no message) | Improved vaccine intentions (convenience framing), no effect on health information seeking or vaccination rates |
| De Anda (2022)<br>*USA* | 680 English or Spanish speakers aged 15+ at COVID testing site | In-person education by community members of same cultural background (vs. no education) | Improved psychological distress, no effect on attitudes towards vaccine or preventative behaviour. |
| Duong (2024)<br>*Vietnam* | 303 college students | News stories framing risk of infection as high or low | Improved preventative behaviour and information sharing intentions (fear or comparative optimism mediated effect), increased, fear, anxiety and anger (high risk framing). |
| Egan (2021)<br>*UK* | 4,099 adults (nationally representative) | Infographics or text-only messages (vs. no information) | Improved knowledge and confidence (infographics) |
| Elliott (2023)<br>*Global* | 295 parents of children under 18 | Plain language message (vs. standard language) | Improved knowledge no effect on behaviour. |
| Felgendreff (2024)<br>*Germany* | 1,810 unvaccinated adults | Infographics (vs. no information) | Improved attitudes towards vaccine, no effect on vaccine intentions. |
| Freeman (2021)<br>*UK* | 18,855 adults (nationally representative) | Persuasive written messages (personal vs. collective benefits, seriousness, safety concerns, or combination of all). | Improved vaccine intentions (personal benefit, safety concerns and combination condition only in strongly hesitant individuals) |
| Frias-Navarro (2021)<br>*Spain, Chile, & Columbia* | 3,362 adults | Facebook messages with 4 moral frames (deontological, utilitarianism, promotion of virtue) | Reduced perceptions about others' behaviour, preventative and information sharing behaviour intentions (virtue frame) |
| Helfers (2023)<br>*Germany* | 588 unvaccinated adults | Infographics debunking misinformation (vs. usual care) | Reduced misinformation beliefs (in medium a priori misinformation group), increased misinformation beliefs(in high a priori misinformation group. No effect on vaccine intentions. |
| Hing (2022)<br>*Malaysia* | 5,788 unvaccinated adults | Persuasive written messages (Descriptive norms; negative vs. positive attributes; risk vs. no persuasion) | Improved information sharing intention with those with pre-existing condition (descriptive norms and positive attributes framing). No effect on vaccine intention |
| Igartua (2023)<br>*Spain* | 278 uninfected adults aged 18-28 | Messages including fictional testimonial (low vs high severity frame; protagonist vs protagonist father) | Improved preventative behaviour intentions ( severe framing via indirect mediation by cognitive elaboration and reduced reactance). Increased perceived personal risk (severe framing via indirect mediation by cognitive elaboration). |
| Koban (2023)<br>*USA* | 371 unvaccinated adults | Professional-led Facebook group (vs. usual care) | Improved vaccine confidence and intentions (mediated via trust) |
| Kopelowicz (2023)<br>*USA* | 100 Latinx, Spanish speaking adults with schizophrenia | Animated, story-based video in Spanish (vs. unrelated animation) | Improved vaccine intentions, no effect on knowledge, other attitudes or behaviours. |
| Legate (2022)<br>*89 countries* | 25,718 adults | Persuasive written messages (autonomy supportive, pressuring or no message) | No effect on preventative behaviour intentions (autonomy or pressuring frames); decreased defiance (autonomy frame); pressuring message group had more controlled motivation than no-message |
| Li (2023)<br>*China* | 981 unvaccinated adults | Persuasive written messages (gain and loss frames vs. usual care) | Improved vaccine intentions (both gain & loss framing). |
| Li (2024)<br>*Australia* | 226 adults | Health literacy sensitive infographic vs. usual care vs. no information | Improved knowledge (infographic); no effect on preventative behaviour intentions. |
| Lungu (2023)<br>*Norway* | 1,192 ambulance charity members | Videos including calls to actions, focus on various health issues and expert vs. non expert sources | Improved knowledge (call to action); Improved trust (expert source & call to action); Improved preventative behaviour intentions (non-expert source) |
| Massey (2022)<br>*USA* | 1,004 adults smokers | Persuasive written messages (COVID only risk vs. smoking risk vs. combined risk vs. unrelated message) | No effect on COVID-19 perceptions; No effect on misinformation beliefs; Increased perceived smoking severity (combined framing); Improved preventative behaviour intentions (combined framing). |
| Merle (2024)<br>*USA* | 350 adults | Text vs. video vs. infographic | No effect/difference between groups on preventative behaviour intentions. |
| Motta (2021)<br>*USA* | 7,064 adults (nationally representative) | Persuasive written messages (personal vs. collective vs. economic benefit; expert vs. non-expert source; presence or absence of prebunking) | Improved vaccine intentions (personal & collective health framing) |
| Okuhara (2020)<br>*Japan* | 1,980 uninfected adults | Persuasive written messages (Expert, physician, authority figure, patient, resident of outbreak area vs. unrelated information) | Improved preventative behaviour intentions, Increased perceived risk, Increased response efficacy, Increased self-efficacy (physician source) |
| Okuhara (2022)<br>*Japan* | 969 adults with children under 11 | Persuasive written messages (kin care vs. avoidance vs. unrelated information) | Improved vaccine attitudes and intentions (kin care & avoidance) |
| Reddinger (2022)<br>*USA* | 2621 unvaccinated adults | Persuasive written messages (tailored to identity: Black, Latinx, conservative, religious, or being parent) | No effect/difference on vaccine hesitancy |
| Ryan (2023)<br>*USA* | 89 senior food assistance program participants | Flyer and calls from health coach (vs. usual care) | No effect on knowledge; Improved preventative behaviours; Increased vaccination rates |
| Saling (2024)<br>*Global* | 213 adults | Exposure to accuracy-sensitisation prime (vs. no prime) | Improved information sharing intentions (primed group) |
| Sayfi (2024)<br>*Global* | 488 adults aged over 21 | Plain language message from WHO or CDC vs. standard language | Improved knowledge, information sharing intentions, preventative behaviour intentions (plain language) |
| Smith (2024)<br>*South Africa* | 81 adult COVID-19 cases and contacts aged over 12 | In person community worker led education (vs. usual care) | No effect on household transmission. |
| Thaker (2021)<br>*New Zealand* | 1,083 adults (nationally representative) | Exposure to vaccine misinformation, hesitancy or factual information. | Reduced believability of misinformation, reduced vaccine and information sharing intentions (misinformation & hesitancy frames) |
| Thunstrom (2021)<br>*USA* | 3,133 adults (nationally representative) | Persuasive written messages (high vs. low probability infection; high vs. low risk of fatality; CDC & White House source vs. CDC only source) | Reduced perceived risk (low risk frame); Reduced vaccine intentions (low-risk & inconsistent information frames) |
| Torres (2021)<br>*USA* | 18,223 Black or White non-college educated adults | Video message from Black or White physician (vs. unrelated video) | Improved knowledge and increased information sharing and willingness to pay for mask. No effect on behavioural intentions. |
| van Empelen (2022)<br>*Netherlands* | 424-593 adults | Volitional help sheet, behavioural journalism, empathy induction (vs. no information) | Increased perceived disease severity, susceptibility and preventive behaviour intentions (help sheet format); Increased perceived vulnerability (empathy induction format) |
| Vandormael (2021)<br>*USA, Mexico, UK, Germany, Spain* | 14,482 adults | Animation video (vs. unrelated video or no video) | Improved knowledge and preventive behaviour intentions (animation) |
| Vivion (2022)<br>*Canada* | 2,500 unvaccinated adults aged over 50 | Inoculation message (vs. unrelated message) | Improved vaccine intentions (inoculation) but no effect on attitudes. |
| Yildirim (2020)<br>*USA* | 2,617 adults | Persuasive written messages (“equal pandemic” vs. “elderly and medical conditions” vs. “class inequality”) | Reduced perceived risk and increased support for economy (elderly inequality frame) |
| Yu (2022)<br>*China* | 296 vaccinated adults | Educational leaflet (vs. none) | Improved vaccine knowledge |
| Yuan (2022)<br>*USA* | 702 unvaccinated adults | Video messages framed on individual vs. community vs. country-level benefit | Improved attitudes towards mandates (individual frame); Improved vaccine intentions (individual frame) |
| Yuen (2023)<br>*Hong Kong* | 1,072 adults | Persuasive written messages (neutral government announcement vs. health expert supporting government vs. health expert disagreeing with government) | Improved attitudes towards some prevention behaviours (consensus frame); increased trust in health experts (contradiction frame) |
| Zhang (2024)<br>*China* | 2,526 primary school children in 11 schools | Health education lecture and videos (vs. usual care) | Improved knowledge but no effect on vaccination rates. |
| Zhao (2024)<br>*China* | 371 adults | Persuasive written messages (certain vs. uncertain solution, certain vs. uncertain solution) | Increased perceived severity and improved information seeking intentions (uncertain solution frame); Increased misinformation susceptibility (uncertain solution & high threat frame) |

#### Intervention aims

Common aims were to provide information on how to influence public perception/attitudes about COVID-19 and preventative measures against COVID-19,^16, 20, 22, 24, 27, 29, 33–35, 37, 39–53^ and to enhance COVID-19 related health behaviours or intentions to engage in these.^13–17, 20–23, 25–34, 36–39, 41–48, 51, 53–59^ All studies focussed on the knowledge translation activity of disseminating scientific information to the public.

#### Intervention Format

Of the 48 studies, 33 were conducted online ^14–22, 24, 25, 28, 30–33, 38, 40, 41, 43, 45–49, 51–53, 55–58, 60^ and 15 were conducted in applied settings.^13, 23, 26, 27, 29, 34–37, 39, 42, 44, 50, 54, 59^ The types of interventions examined by the included studies were primarily public health communication, ^13–16, 19–21, 23, 25, 28, 31–33, 38, 43, 45–49, 51–58, 60^ educational interventions ^17, 18, 26, 29, 34–37, 39–42, 44, 50, 59^ or both.^22, 24, 27, 30^ Many interventions were formatted as written text messages ^13, 15, 16, 19–23, 25, 30, 33, 43, 52, 54, 55, 57, 60^. Some interventions used visual formats like infographics ^22, 24, 25, 27, 28, 56^ or videos.^17, 24, 39–41, 44, 45, 47, 48, 52, 56^ Some interventions were messages styled as news stimuli ^38,46, 53^ or social media messages.^14, 19, 43^

Interventions in applied settings included in-person education programs and physical educational resources,^37, 42, 59^, as well as online educational programs, ^34, 36^ including professional led education programs delivered via social media groups.^26, 29, 35^

#### Risk of bias

Of the 48 included studies, 42 had an overall low risk of bias and 6 had some concerns. The most common sources of bias were inadequate randomisation procedures (n = 9) and measurement of outcome (blinding) (n = 5). Full details in Online supplemental file 3.

#### Meta analysis

There were papers that measured similar outcomes for a meta-analysis, however, as interventions were considered too qualitatively different, the final meta-analysis were not valid for meaningful interpretation. This is reinforced by the measures for heterogeneity in the random-effects models for knowledge outcomes (N=8 ;*I*^2^=0.92) and the vaccine intentional outcome (N=5; *I*^2^=0.91). Forest plots in Online supplemental file 4 and 5. Additional methods for assessing heterogeneity and sensitivity analyses were not further explored as final meta-analysis was not valid for meaningful interpretation and any subgroup analyses were too small.

### 2. Psychological outcomes

A total of 40 studies reported on psychological outcomes, with 30 (75%) reporting a positive effect of the intervention.

### Knowledge

Of the 15 studies that measured knowledge outcomes, eight focused on disease (COVID-19) knowledge and six on vaccine knowledge, with other studies focussing on COVID-19 testing, masking and other mandate-related knowledge.

Most of the interventions effectively improved knowledge (13 of the 15 studies). Nearly all (7 of the 8) video or animation-based interventions aiming to improve knowledge were effective ^17, 24, 39–41, 45, 47^ with the exclusion of one animated, story-based video intervention.^44^ Other effective formats included infographics,^22, 25^ and messages that used plain language,^13, 16^ including calls to action ^45^ and virtual social media-based coaching by pharmacists and physicians.^35^ An intervention matching race of physician to the audience had positive effects on knowledge, especially in Black participants in USA.^47^ An intervention using an in-person bilingual health coach, however, had no effect on knowledge.^37^

#### Attitudes and beliefs

Of the 29 studies that measured attitudes and beliefs, 13 focussed on attitudes towards vaccines (i.e. hesitancy, confidence), 10 towards disease (i.e. perceived risk and severity of COVID-19), and 11 towards other prevention behaviours (i.e. effectiveness of protective behaviours, necessity of mandates). A further five studies focussed on self-efficacy, five on misinformation beliefs and three on trust in science. Nineteen of the studies found the interventions had a positive effect, six found no effect and four found negative effects. Negative effects occurred in the studies that tested different messaging framings, and included reducing perceived risk of disease,^21, 49^ reducing perceptions about others adhering to preventative behaviours ^14^ and leading to increased controlled motivation.^15^ For example, Yildirm et al. ^49^ found framing messages around the difficulties elderly and individuals with medical conditions faced in the pandemic led to increased support for saving economy instead of saving lives, in comparison to the group that received “equal pandemic” framing which emphasised how the outbreak affected everyone regardless of background.

Four interventions were administered via online formats including social media or online course and SMS formats ^26, 29, 35, 36^ and were all effective in improving attitudes and beliefs. For example, three studies on social media coaching interventions found improved attitudes towards vaccines and increased personal responsibility,^26, 29, 35^ and a study using an online educational course and SMS reminders improved grandparent’s attitudes towards getting a COVID-19 booster dose.^36^ Six out of seven studies with videos and infographics improved attitudes; two improved attitudes towards vaccines,^27, 40^ two towards disease and its prevention,^41, 48^ and two reduced misinformation beliefs or increased trust in science.^28, 45^

Six studies used professional or expert sources such as health professionals as part of the intervention, with all reporting positive results on attitudes and beliefs.^26, 29, 35, 45, 52, 58^ In one study, Yuen et al. ^52^ tested messages featuring health experts who either supported or opposed mandated COVID-19 measures. Expert agreement increased support for banning public assembly, however it had no effect on support for contact tracing mobile apps or restriction testing. This study also found that when participants were shown health experts disagreeing with measures, their trust in health professionals increased.

Tailoring messaging to the audience and using different framings/persuasive messaging techniques had variable results – nine out of seventeen studies reported positive effects, four found no effect in,^19, 32, 33, 46^ and four found negative effects.^14, 15, 21, 49^ Bokemper et al. ^20^ found that framing preventative health messages around the “recklessness” of not social distancing and the importance of collective action improved preventative health attitudes. Two studies ^23, 51^ found that framing messages around individual or personal benefits improved attitudes towards vaccines and mandates, particularly in the strongly vaccine hesitant group. ^23^ Two studies that varied descriptions of disease severity in messaging found that framing severity as high improved attitudes towards disease,^43^ whereas low risk framing had the opposite effect, leading to reduced perceived risk of disease.^47^ Messages with social comparison to a “morally ideal” person reduced intentions to carry out public health behaviours, and decreased perceptions of others’ likelihood of engaging in these same behaviours.^14^ Messages highlighting unequal impact of COVID-19 on the elderly and individuals with medical conditions reduced perceived risk of disease, and instead favoured saving economy over saving lives.^49^

One study testing messaging with combined cognitive claims (e.g. statements about scientific results) and normative claims (e.g. what should be done given those scientific results) with messaging with cognitive claim only found neither improved trust in science.^19^ An intervention for smokers found combining COVID-19 and smoking messages (e.g. underscoring how smoking makes worse), was also ineffective for improving attitudes towards disease or misinformation.^46^ Tailoring the message based on audience identity (e.g. ethnicity, religion, political stance and being parent) had no effect on vaccine hesitance.^32^

The five studies focussing on misinformation beliefs and vaccine attitudes also had mixed results, with only two showing a positive effect. Debunking misinformation was effective in people who held medium levels of a priori misinformation beliefs,^28^ and exposure to outright misinformation and vaccine hesitancy messages led to lower believability of misinformation.^60^ However, one study found that participants exposed to vaccine hesitancy messaging were more likely to believe it compared to the group exposed to outright misinformation messaging.^60^ Two other studies ^33, 46^ found no effect on misinformation beliefs of interventions that inoculated or pre-bunked misinformation, or combined risk framing of COVID-19 with smoking. Another study found messages that highlighted uncertainty and high threat of disease were found to increase misinformation susceptibility. ^53^

#### Emotions

Of the three studies that focussed on emotion, two ^15, 42^ led to reduced negative emotions; for example, De Anada et al. ^42^ reduced psychological distress in their community-led in-person intervention, and Legate et al. ^15^ reduced defiance by using autonomy-supporting messaging. Another intervention ^38^ tested different disease risk framings and found the high risk framing led to increased fear, anxiety and anger.

### 3. Behavioural outcomes

A total of 40 studies reported on behavioural outcomes, with 28 (70%) reporting positive effects of the intervention. Most of the studies focussed on behavioural intentions; only nine studies reported actual behaviours via self-report methods. Twenty studies focussed on vaccination related behaviours, 19 on other preventative health behaviours and 12 on health information seeking/sharing behaviours.

#### Vaccination

A total of 20 studies measured effects on vaccination (self-reported rates and intentions). Three studies ^21, 30, 57^ were conducted before vaccines were available to the public, measuring hypothetical intentions. Twelve studies found a positive effect of the interventions, two found negative effects, and six found no effects. All four studies which involved online professional-led groups improved vaccination self-reported rates and/or intentions.^26, 29, 35, 36^ An intervention that used bilingual community workers with regular check-ups also led to an increase in vaccination rates.^37^ Of the three video interventions targeting vaccination, ^39, 41, 44^ only one improved vaccine intentions.^44^ Whilst there are no notable difference between the three studies that could have caused the disparity, the effective intervention was a Spanish animation, whilst the unsuccessful interventions were an English animation intervention ^41^ and a school-based video program in China.^39^

The two studies that used infographics had no effect on vaccine intentions.^27, 28^ Manipulating the framing of the message had mixed effects on intentions. Framing vaccines as “free, available and easy to obtain”,^54^ or focussing on personal/individual benefits ^23, 51, 57^ increased intentions. Framing the importance of vaccination on kin care (caring for family) or disease avoidance also increased intentions,^31^ as did both gain and loss framed messages.^30^ Using descriptive norms had no effect ^55^ and framing risk of disease as low reduced intentions.^21^

The three studies that focussed on misinformation and vaccination intentions also had mixed results; pre-bunking improved intentions,^33^ debunking had no effect,^28^ and exposure to misinformation and hesitancy information reduced intentions.^60^

#### Other preventative behaviours

A total of 19 studies measured effects on other preventative behaviours (14 reported on intentions only, three reported on actual behaviour and two reported on both actual behaviour and intentions) like social distancing, mask wearing and hand washing. Twelve of these had a positive effect, with six studies having no effect and one having negative effects on preventative behaviours.

Studies that focussed on plain language and health literacy sensitive formats had mixed results, with three of five studies reporting positive effects. Plain language formats ^16^ and volitional help sheets ^48^ were found to improve behavioural intention, whilst infographics studies had mixed results.^22, 25, 56^ One study ^22^ found that infographics improved intentions of mask wearing compared to official government text information, whereas two other studies found infographics had no impact on intention to self-isolate in hypothetical scenarios ^25^ or practice handwashing guidelines.^56^

Video-based interventions had mixed results, with two of five studies reporting positive effects. For example, an animation intervention improved intentions to wash dishes to mitigate COVID-19 spread ^17^, yet video interventions in other studies did not improve behavioural intentions or self-reported behaviours.^44, 56^

Whilst featuring credible sources typically improved preventative behaviours and intentions,^34, 58^ non-expert sources improved intentions in comparison to expert sources in another study.^45^ Matching the race of the physician delivering the information to the intended audience had no effect on self-reported safety behaviours.^47^ Similarly, there were mixed results with interventions that utilised community workers; bilingual health coach check-ups improved COVID-19 mitigation behaviours compared to control in one study,^37^ but had no effect on self-reported behaviour in another.^42^

Persuasive messaging techniques showed mixed effects on preventative behaviours, with five of seven studies reporting positive outcomes. For example, combining COVID-19 and smoking risk messaging increased mask wearing intentions,^46^ and framing failure to socially distance as reckless and emphasising collective action improved social distancing intentions.^20^ High/severe risk framings were also effective in increasing intention to adopt preventative measures, mediated either by cognitive elaboration and reduced reactance,^43^ or by fear or comparative optimism.^38^ In contrast, framing messages around their ethical virtue reduced intentions to wash hands and stay home,^14^ and messages that were either controlling or autonomy-supportive had no effect on social distancing intentions.^15^

#### Health information sharing and seeking

Interventions aiming to improve information-seeking or sharing had mixed results; of the ten studies, only five found a positive effect and no consistent intervention type emerged as clearly effective. For example, of the two studies focusing on plain language formats, one found no effect,^13^ whilst the other found positive effects.^16^ The one video intervention targeting sharing intentions also found no effect.^41^ Interventions testing different framings had variable results; matching race to audience ^47^ and frames which highlighted uncertainty ^53^ improved information-seeking behaviour and intentions, whilst moral virtue framings reduced intentions.^14^ One study which tested different norms and freedom/gain framings found no effects of any framing on information sharing intentions,^54^ whilst another found descriptive norms improved intention to share information to those with pre-existing conditions.^55^ Likewise, of the two studies focussing on misinformation and sharing intentions, one found priming participants to consider information accuracy before showing misinformation had a positive effect on information sharing intentions,^18^ whilst another found exposure to misinformation and hesitancy had negative effects on information sharing intentions.^60^

#### Behaviour change techniques

Figure 2 provides details of behaviour changes techniques (BCTs)^9^ included in the intervention groups for studies that measured behavioural outcomes and provided sufficient content detail in English. The most common BCTs used in the studies’ interventions were ‘Information about health consequences (5.1.)’, ‘Credible source (9.1.)’, ‘Instruction on how to perform a behaviour (4.1.)’, ‘Salience of consequences (5.2)’ and ‘Information about social and environmental consequences (5.3.)’. The study designs compared different complex interventions and did not use established taxonomies to describe intervention differences, so effects could not clearly be attributed to specific BCTs.

**Figure 2.**
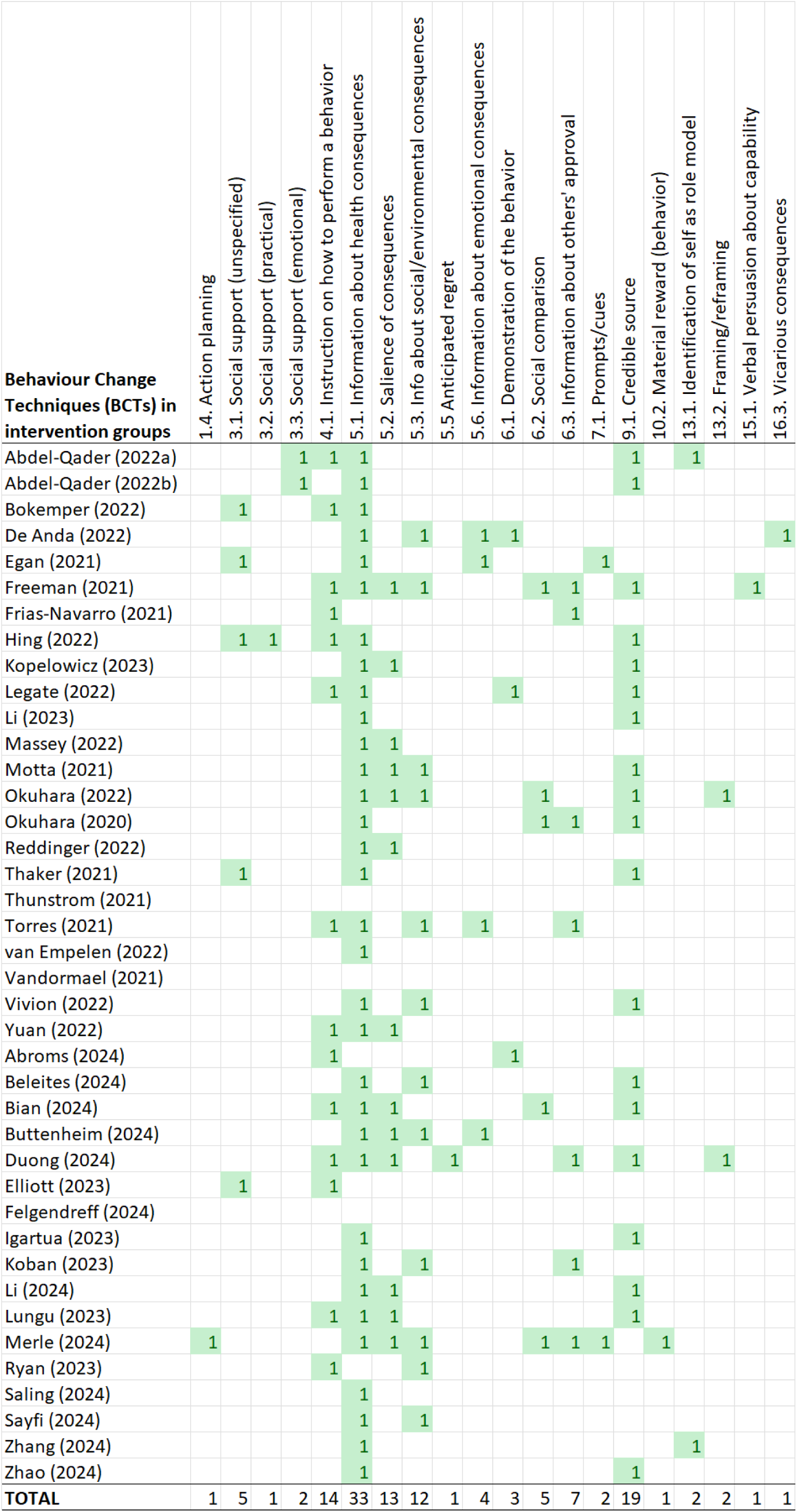
Behaviour Change Techniques used in the intervention groups of studies that measured behavioural outcomes

### 4. Health outcomes

Only one study focussed on actual health outcomes (infection/disease transmission).^59^ Smith et al. ^59^ evaluated the effect of a resource kit delivered in person by community health workers on COVID-19 transmission. It found that household transmission mitigation was ineffective, since participants had significant exposure to infection prior to the study.

## DISCUSSION

This review identified 48 RCTs on community-focussed COVID-19 science and knowledge translation strategies. The studies were extremely heterogenous, but common features of effective interventions included credible sources and interactive audiovisual formats. Effects of tailoring information to different population groups were less clear, and exposure to misinformation or framing disease as low risk had negative impacts. A wide range of BCTs were used across the studies, but the studies were not designed in a way that could identify the specific effect of individual BCTs, or comparisons between BCTs.

The findings align with more specific systematic reviews related to COVID-19, including vaccine uptake,^61–64^ physical distancing or other behaviour changes,^65–67^ and strategies to counter misinformation.^68–70^ Other reviews also reported that studies were highly heterogenous,^62^ with few measuring actual behaviour,^61, 62^ highlighting a major limitation to the science translation evidence produced during COVID-19. However, credible sources such as health experts, physicians and other health professionals appear to consistently enhance the effectiveness of COVID-19 communication interventions.^64, 66^. Reviews focused on COVID-19 vaccine uptake and willingness also support the value of interactive and digital/online formats.^62, 63^ Notably, Kuwahara et al.^67^ found that digital COVID-19 messages delivered by health professionals which combined credible sources with digital delivery can promote behavioural change *and* reduce infection rates. Previous studies of specific communication strategies such as message framing have been mixed.^63, 64^ Communicating with overly certain messaging, rather than acknowledging uncertainty around vaccine efficacy or risks, has been shown to backfire in other reviews.^62, 70^ A review by Epton et al.^65^ found two of the most commonly reported BCTs in this study, ‘information about health consequences’ and ‘highlighting salience of health consequences’, were effective for promoting physical distancing during pandemics. Reviews on interventions addressing misinformation similarly reported mixed effectiveness for both pre-bunking and debunking strategies.^68–70^. However, the review by Smith et al.^69^ found that accuracy sensitisation primes were effective in reducing misinformation beliefs.

This review highlights the lack of theory-based study design to isolate the effect of specific BCTs, and a need for standardised measures to evaluate science and knowledge translation across studies to enable comparison. The use of pragmatic study designs with complex multicomponent interventions is understandable given the emergency context. However, if a common framework had been used, the 48 studies could have offered clearer insights into which techniques were most effective in different contexts, to inform future pandemic preparedness. The COVID-19 pandemic provided an opportunity to advance knowledge in behavioural science, however coordinated efforts were missed in the urgency of responding rapidly to an unfolding emergency. Whilst some efforts to standardise behavioural science exist (e.g. the BCT taxonomy used in this review), an internationally accepted, core outcome set for emergency pandemic contexts could improve the uptake of standardised approaches to study design and evaluation. Platforms developed during COVID-19 could be adapted for this (e.g. [Cochrane’s COVID-19 methods initiatives](https://www.cochranelibrary.com/cdsr/doi/10.1002/14651858.CD202002/full)^71^), alongside broader approaches (e.g. [COSMIN](https://www.cosmin.nl/)^72^), to assist researchers to clearly identify and test potentially effective BCTs for use in different settings. Knowledge translation platforms that synthesise evidence from multiple sources could further promote public use, with user-friendly formats and plain language to bridge the gap between scientific evidence and the public.^73^

The review followed a rigorous, pre-registered protocol based on PRISMA guidelines, and included a theory-based behavioural science taxonomy to compare 48 studies in a standardised way. Due to the heterogeneity of the studies, meta-analyses were inappropriate for this data. The complex interventions and study designs made it difficult to isolate the effects of specific intervention components across studies. Inconsistencies in author use of science and knowledge translation terms may have affected the comprehensiveness of the search. Finally, most studies measured behavioural intentions rather than actual behaviour, and actual behaviour outcomes were self-reported.

### Conclusion

This review found a wide range of science and knowledge translation strategies reported to be effective during the COVID-19 pandemic. Better use of behavioural science taxonomies and core outcome sets could help researchers advance the field further during future health emergencies, by isolating the most effective techniques to use in different contexts.

## Data Availability

No new data were generated. The protocol for this systematic review is registered in PROSPERO (CRD42023446093). Search strategies and data extraction tables are available in the supplementary materials.

## Funding

Carissa Bonner was supported by a NHMRC fellowship. A related rapid scoping review was funded by the World Health Organization.

## Competing interest

We declare no interests.

## Author statement

CB, KM and JB conceived and designed the study. MT, MF, EF, RK and CB extracted and analysed the data and drafted the manuscript. All authors reviewed and contributed to the final manuscript.

## Acknowledgements

We thank the participants of the ‘WHO Informal Consultation with Experts for Science and Knowledge Translation in Health Emergencies’, for feedback on preliminary scoping review results (Istanbul, Türkiye; February 2023); and members of the WHO Knowledge Translation in Health Emergencies Community, for feedback on preliminary systematic review results (online community of practice meeting on the WHO Hive digital collaboration platform; September 2024).

## Data availability

Data available upon reasonable request. Search strategy and data extraction form available in supplementary material.

## Notes

### Competing Interest Statement

The authors have declared no competing interest.

### Summary of Updates

Minor typographical edits made in title. Further details given in abstract.

