## Supplementary material for "Science and knowledge translation strategies to the public during health emergencies: systematic review of RCTs": online supplemental file

### Online supplemental file 1. Search strategy

| **Database** | **Search Strategy 26-11-2024** | **Limiters** |
| --- | --- | --- |
| PubMed | (“Science Translation” OR “Science Education” OR  “Science Literacy” OR “Science Communication” OR  “Knowledge Translation” OR “Health Literacy” OR “Health Education” OR “Health Communication” OR “Public engagement” OR “Knowledge exchange”) AND (Community OR Citizen OR “Social Media” OR Public) AND (COVID-19) | Randomised Controlled Trials, Clinical Trial |
| Scopus | (“Science Translation” OR “Science Education” OR  “Science Literacy” OR “Science Communication” OR  “Knowledge Translation” OR “Health Literacy” OR “Health Education” OR “Health Communication” OR “Public engagement” OR “Knowledge exchange”) AND (Community OR Citizen OR “Social Media”) AND (COVID-19) AND ("Randomi?ed Controlled Trial*" OR "RCT" OR "Randomi?ed" OR “Clinical Trial”) | Article |
| CINAHL | (“Science Translation” OR “Science Education” OR  “Science Literacy” OR “Science Communication” OR  “Knowledge Translation” OR “Health Literacy” OR “Health Education” OR “Health Communication” OR “Public engagement” OR “Knowledge exchange”) AND (Community OR Citizen OR “Social Media” or Public) AND (COVID-19) AND ((MH "Randomized Controlled Trials+") OR (MH "Clinical Trials+") OR (MH "Cochrane Library") OR (MH "Clinical Trial Registry")) | Nil |
| PsycINFO | ("Science Translation" or "Science Education" or "Science Literacy" or "Science Communication" or "Knowledge Translation" or "Health Literacy" or "Health Education" or "Health Communication" or "Public Engagement" or "Knowledge Exchange").mp  (Community or Citizen or "Social Media" or Public).mp.  exp COVID-19/  limit 4 to ("0300 clinical trial" or "0430 followup study" or "0450 longitudinal study" or "0451 prospective study") | Nil |

### Online supplemental file 2. Data extraction form

All data extracted from Articles should be copied and pasted text that meets the following descriptions

| **Data** | **Description** |
| --- | --- |
| *Characteristics* | |
| Title | Title of the journal article |
| First Author | The first author’s surname |
| Publication Year | The year in which the journal article was published |
| Study Design | The design used in the study |
| Country | The current the study was conducted in |
| COVID Context | Outbreak level, specific variant or anything about restrictions when the study was conducted (if reported in the paper) |
| Year Data was Collected | When was the data collected in the study |
| Author Defined KT | Yes/Partially (not explicitly referencing KT)/ No |
| KT Definition | How did the authors defined KT (if they did)  OR How do reviewers conceptualise how KT was defined by this study. |
| Knowledge Translation (KT) Code | Generation / Dissemination |
| *Sample Characteristics* | |
| Ethnicity/Race | What race/ethnically statistics are provided by authors that describe their studied sample |
| Age | As above, for age |
| Sample Size | As above for sample size |
| *Intervention Information* | |
| Intervention | Describe what the intervention is in detail |
| Intervention Code | Single word/phrase reviewer summary of the intervention |
| Comparator | Describe what comparators. |
| Comparator Code | Single word/phrase reviewer summary of the comparator |
| **Outcome information** | |
| Implementation | Uptake, reach, cost |
| Psychological | Knowledge |
| Behavioural | Intention, Self-report |
| Health Outcomes | Incidence of COVID |
| For each outcome measured:   1. Add two columns for the specific outcomes measured (one for the description of what the outcome is and the other for the measures used (point estimate and 95% CI interval) 2. Only add two new columns if there is a new outcome. 3. Main outcome data that is extracted should be active-to-control comparisons. 4. If there is an active-to-active comparison, the data extracted should make clear that the data refers to this   Alternative, tables can be extracted | |

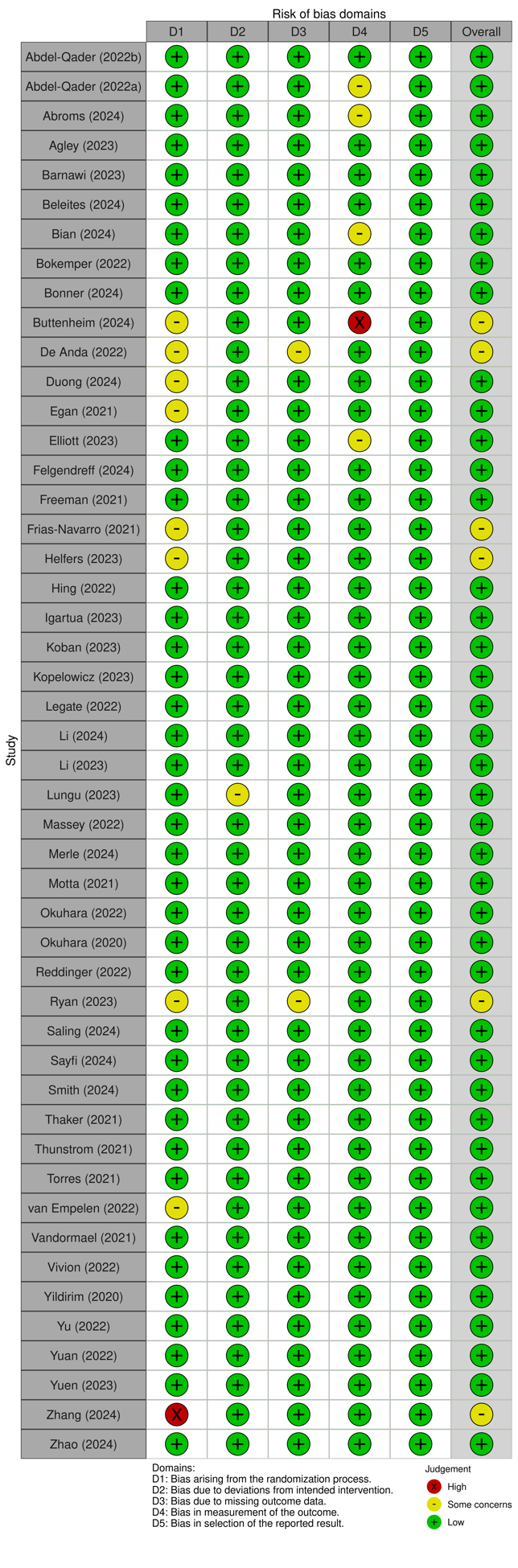

### Online supplemental file 3. Risk-of-bias visualisation

### Online supplemental file 4. Forest plot for meta-analysis for knowledge outcomes

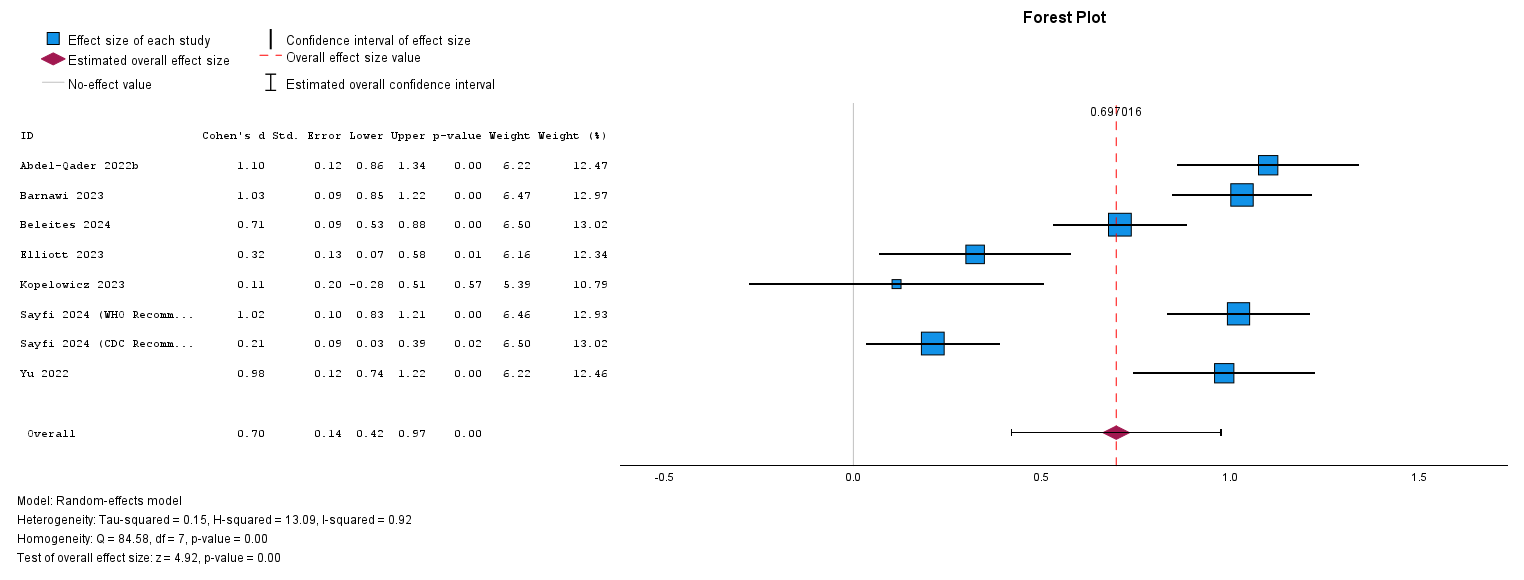

### Online supplemental file 5. Forest plot for meta-analysis for vaccine intentional outcomes

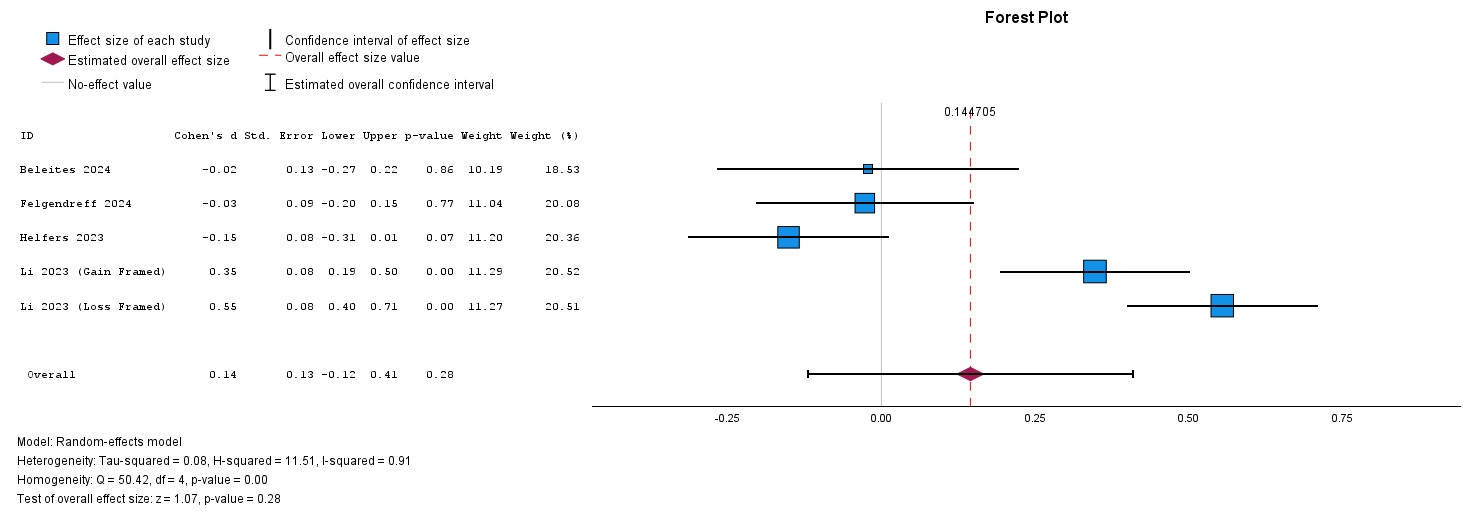

### Online supplemental file 6. Detailed summary of included studies

| **Study** | **Design** | **Sample** | **Intervention** | **Outcomes** | **Key findings** |
| --- | --- | --- | --- | --- | --- |
| Abdel-Qader (2022a)  *Jordan* | RCT with 2 groups - intervention and control. | 295 Jordanian adults who were disbelieving towards COVID-19, unwilling to receive COVID-19 vaccinations, and usually not engaging in preventative behaviours for COVID-19. Individuals who had already been infected with COVID-19, had received a COVID-19 vaccine were ineligible to participate. | 12 weekly pharmacist-delivered virtual coaching sessions via Zoom. Topics included COVID transmission pathways and precautionary measures to block these, severity and long-term effects of COVID-19, vaccines available, each individual’s role in the pandemic control. Pharmacists also provided emotional support. | ***Behavioural:*** Preventive behaviours measured (face masks, using hand sanitisers, disinfecting surfaces, avoiding touching T-zone, maintaining physical distance, using disposable tissues, avoiding crowds and others and sharing objects, and self-isolation) | - Intervention produced a significant increase in COVID-19 preventative behaviours compared to passive control. |
| Abdel-Qader (2022b)  *Jordan* | RCT with 2 groups - intervention and control. | 305 adult Jordanian residents who were hesitant or resistant to COVID-19 vaccine. Those without a Facebook account, who were willing to take the vaccine or had taken one or more doses of the COVID-19 vaccine, or who currently had or had already recovered from COVID-19 were excluded. | Pharmacist & physician-delivered virtual coaching sessions – 16 sessions delivered through Facebook live over a 2 month period. | ***Psychological:*** Knowledge of, attitude towards and beliefs about COVID-19 vaccine, COVID-19 vaccine hesitancy  ***Behavioural*:** Getting COVID-19 vaccine | - Intervention significantly reduced COVID-19 vaccine hesitancy and resistance both immediately post-coaching and at 1-month follow up. - Knowledge of, attitude towards and beliefs about COVID-19 vaccine all significantly changed favourably after intervention - Vaccination rates in coaching group increased a month after coaching whereas control group did not |
| Abroms (2024)  *USA* | RCT with 2 groups - intervention and control. | 478 adults residing in the USA, who had not received a single dose of the COVID-19 vaccine, and were daily Facebook users. | A Facebook group where moderators posted two informational posts per day for 4 weeks and engaged in relationship-building interactions with group members. | ***Psychological:*** COVID-19 vaccine confidence, COVID-19 vaccine complacency, COVID-19 vaccine necessity, general vaccine confidence and responsibility to vaccinate.  ***Behavioural:*** Getting the COVID-19 vaccination and intention to vaccinate. | - No differences were found in vaccination rates. - Intervention participants were more likely to show improvements in their COVID-19 vaccination intentions (vs. stay same or decline) compared with control (p = .03). - They also improved more in their intentions to encourage others to vaccinate for COVID-19. - There were no differences in COVID-19 vaccine confidence or intentions between groups. - General vaccine and responsibility to vaccinate were higher in the intervention compared with control. |
| Agley (2023)  *USA* | 2-armed, post-test only randomised controlled experiment | Nationally representative US sample by cross sections of age, sex, and race/ethnicity (n=1526). | Social media messaging with cognitive claims (e.g. statements about scientific results) and normative claims (e.g. what should be done given those scientific results). | ***Psychological:*** Trust in science | - Hypotheses that single exposures to normative language can reduce perceptions of trust or credibility in science or scientists for all people were not supported - Secondary analyses indicate the possibility that political orientation may differentially mediate the effect of normative and cognitive language from scientists on people’s perceptions. |
| Barnawi (2023)  *Saudi Arabia* | 3-armed, post-test only randomised controlled experiment | Saudi citizens who could read and communication in Arabic or English and were aged 18 or above (n=508). | Educational video interventions containing clips of the types, components, and mechanisms of approved COVID-19 vaccines, disclaimers of COVID-19 vaccine related rumors based on scientific evidence. | ***Psychological***: Knowledge about vaccination; concern around COVID-19 vaccination | - The experimental group showed a significantly lower average level of concern about the COVID-19 vaccination than the control group. - The experimental group showed a significantly higher average level of knowledge about the COVID-19 vaccination than the control group. |
| Beleites (2024)  *USA* | Online RCT with three groups – intervention, attention-placebo control and inactive control. | 792 citizens or permanent residents of the United States (US), who were aged at least 18 years old and native English speakers. | A collage of three Short, Animated, Story-based (SAS) videos, each emphasizing the importance of COVID-19 vaccination. | ***Implementation***: Choice to watch the video post-trial (if in the non-intervention groups).  ***Psychological:*** Vaccine-related knowledge, self & disease perceptions.  ***Behavioural:*** Behaviour intentions - to be vaccinated and to obtain and disseminate credible information about the COVID-19 vaccine. | - Participants in the intervention group displayed significantly higher mean knowledge scores compared to both the attention placebo control and control groups. - Intervention did not notably affect behavioural intent. - The intervention video notably improved scores in perceived response efficacy, perceived social norms, and perceived self-efficacy. - Perception of COVID-19 as a significant health threat was a strong predictor for engaging with the post-trial video, whereas right-wing political inclination negatively associated with engagement |
| Bian (2024) *China* | RCT with 2 groups - intervention and control. | 202 Chinese college students aged 16 years or above, and their grandparents aged 60 years or above. | An online health education program followed by 14 daily SMS reminders to encourage their grandparents to receive a COVID-19 booster dose. Health education materials were delivered to college students through WeChat. | ***Psychological:*** Grandparents’ attitude towards getting a COVID-19 booster dose.  ***Behavioural:*** Uptake of COVID-19 booster vaccination, and intentions to get a COVID-19 booster dose. | - Grandparents in the intervention arm were more likely to receive COVID-19 booster vaccination compared to control cohort - Grandparents in the intervention arm also had greater attitude change and intention change to receive a COVID-19 booster dose. |
| Bokemper (2022) *USA* | 4-armed, post-test only randomised controlled online experiments. | A nationally representative sample of American adults from YouGov survey firm – experiment 1 (n= 3184) and experiment 2 (n= 6079). | Different types of messages that included different aspects of persuasive written messages including social norms, self-interest vs. prosocial concern for social distancing, individual vs. collective action, and values reframing. | ***Psychological:*** Beliefs about social distancing being important for your health and others people’s health and whether an individual would feel guilty for not practicing social distancing  ***Behavioural:*** Intention to social distance. | - A message that reframes not social distancing as recklessness rather than bravery and a message that highlights the need for everyone to take action to protect one another are the most effective at increasing positive beliefs and intentions related to social distancing. |
| Bonner (2024) *Australia* | Online RCT with four groups – 3 intervention groups and one control. | Australian sample of 2005 adults, nationally representative based on age, gender, location, and education. | Three different versions of the risk formats available in the CoRiCal tool Standard CoRiCal: website version with outcomes presented in a series of bar graphs. X per million video: 2 min animation to explain key messages in each bar graph, using probability format   1 in X chance video: 2 min animation to explain key messages in each bar graph, using chance format | ***Psychological:*** Knowledge of COVID-19 and vaccination. | - Both animations increased knowledge compared to standard government information (control). |
| Buttenheim (2024) *South Africa* | Online RCT with 2 experiments, each with 2 intervention and 1 control group. | Experiment 1, 3510 unvaccinated South African adults. Experiment 2, 3608 unvaccinated South African adults. | Exposure to written messages. Experiment 1: social norm message, or a message highlighting that vaccines were free, available and easy to obtain (convenient). Experiment 2: a message highlighting gaining greater freedoms, or a message highlighting being part of the solution to the pandemic. | ***Behavioural:*** Vaccine intentions, vaccine uptake, requests for information.; | - In Experiment 1 - the convenience message significantly increased vaccine intentions. - Experiment 2 - neither value proposition message increased vaccination intentions. |
| De Anda (2022) *USA* | Clustered RCT, with 2 groups - intervention and waitlist control. Assigned at the event level. | 680 community members who were aged 15 years or older, had proficiency in English or Spanish, and visited one of the project’s 43 SARS-CoV-2 testing sites. | In person health education which combined instructions on how to perform COVID-19 preventative behaviours with social support (practical, unspecified) delivered by community health workers of same cultural background as target population. | ***Psychological:*** Attitudes towards preventive behaviours; Vaccine Attitude, Psychological Distress.  ***Behavioural:*** Risky Health Behaviours, Preventive Behaviours | - Only psychological distress was significantly lower in the intervention compared to the control group, other psychological measures were not different - The intervention had no effect on behaviour. |
| Duong (2024)  *Vietnam* | RCT completed in a lab setting using online stimuli. Two experiments with between subjects design (high and low risk conditions). | College students in a large university in Vietnam, aged 18-25 years. The sample size for each of the two experiments was 303. | News stimuli adapted from real news stories from influential online sites, manipulated to describe high or low risk outcomes of Omicron. Additionally, the last paragraph of the news stories in Experiment 1 recommended people to frequently wash their hands with soap to prevent Omicron infection. The recommendation in Experiment 2 encouraged people to test for Omicron infection after traveling, meeting with infected people, or attending crowded public places. | ***Psychological:*** fear, anxiety, anger, comparative optimism.  ***Behavioural:*** Intentions to share health information, and for Experiment 1: Intentions to wash hands with soap and  Experiment 2: Intention to test for COVID-19 when feeling symptoms. | - Results indicated that viewing high-risk news stories led to higher fear, anxiety, and anger than low-risk news stories. - Fear fully mediated the effect of news exposure on intentions to test for Omicron infection and share health information. - Comparative optimism moderated the indirect effect of news exposure on intentions to wash hands with soap and share information about COVID-19 testing. |
| Egan (2021) *UK* | 3x2x2 post-test only randomised controlled online experiment. | Nationally representative sample of adults residing in the UK (n=4099). | Pictorial representations primarily focused on how to correctly place a face mask from start to finish and text-only information. | ***Psychological:*** Recall was focussed upon verifying a respondent’s understanding of how to correctly use a face covering  ***Behavioural:*** Willingness to wear a face mask in public. | - Infographics and several text interventions improved recall - Willingness to wear a mask was high in all participants - Infographics led to highest confidence in mask wearing - However, participants that read text only guidance from UK Government were less willing to do so compared with those that did not read any stimuli. |
| Elliott (2023) *Global* | Online RCT with 2 arms and a nested qualitative component. | 295 parents from anywhere in the world, aged over 18 years, who were a legal guardian of a child under 18 years, and could read and complete the online questionnaire in English. | Plain language and standard language versions of 2 COVID-19 recommendations specific to child health. | ***Implementation:*** Preference, accessibility, usability, satisfaction.  ***Psychological:*** Understanding of pediatric COVID-19 vaccine guidelines.  ***Behavioural:*** Intention to follow recommendation or share information with others. | - Compared to SLVs, parents preferred PLRs and better understood the recommendation. - No significant differences in behavioural outcomes between groups. |
| Felgendreff (2024) *Germany* | Online RCT with 2 experiments. Study 1: 2 intervention groups + control. Study 2 – 2 x 3 between subjects + 1 control group. | Unvaccinated participants from Germany completed the survey, aged 20 – 69 years old. Study 1 (n= 824) ; Study 2 (n= 986). | Infographics on benefits and risks associated with COVID-19 vaccination. | ***Psychological:*** Perceived risk of getting vaccinated, perceived risk of developing blood clots due to Astra Zeneca vaccine.  ***Behavioural***: Vaccine intentions. | - The infographic decreased the risk perception of the vaccine compared to no infographic, but it did not influence the perceived probability of blood clots due to the AstraZeneca vaccine. - The vaccination intention was not affected by viewing the infographic, nor the presented infection rate, but by risk perceptions, sociodemographic characteristics, confidence in the AstraZeneca vaccine, and preference for alternative vaccines. |
| Freeman (2021) *UK* | RCT with 10 x 3 design - ten information conditions stratified by level of vaccine acceptance. | 18855 UK adults (≥18 years of age), who were quota sampled to be nationally representative for age, gender, region, education level, and ethnicity (based on UK Office for National Statistics population estimates) | Written information on the personal benefits and/or collective benefits of vaccination, and/or the seriousness of the pandemic, and/or safety concerns, or a combination of all of these. | ***Psychological:*** Vaccine hesitancy  ***Behavioural****:* Willingness to be vaccinated. | - Information conditions did not alter COVID-19 vaccine hesitancy in those willing or doubtful to be vaccinated. - In those strongly hesitant, COVID-19 vaccine hesitancy was reduced, in comparison to the control condition, by personal benefit information, directly addressing safety concerns about speed of development, and a combination of all information. - In those strongly hesitant, provision of personal benefit information reduced hesitancy to a greater extent than provision of information on the collective benefit of not personally getting ill or the collective benefit of not transmitting the virus. - Ethnicity and gender were found to moderate information condition outcomes. |
| Frias-Navarro (2021) *Spain, Chile, & Columbia* | Online RCT. 3 intervention groups and control group. | Adults living in Spain (n = 1,122), Chile (n = 1,107), and Colombia (n = 1,433). | Four messages in the format of a Facebook message whose content reflects a moral tradition: Deontological ethics, utilitarianism, and promotion of virtue (and control). | ***Psychological:*** Belief about other's behaviour – probability of hand-washing, staying home, sharing the message, and participating in public gatherings.  ***Behavioural:*** Behavioural intention (washing their hands, participating in public gatherings, avoiding all social contact, and sharing the Facebook message they received.) | - The message referring to ethical virtue obtained statistically significant lower scores on the preventative behaviour and information sharing intentions. - The ethical virtue message also had similar negative effect on beliefs about others’ behaviour (only on other people washing their hands, staying at home, and sharing the public health message) |
| Helfers (2023) *Germany* | Online RCT. Two groups – intervention and control. | Adults recruited online (n = 588). Sample was 83% female. Exclusion criteria included being already vaccination against COVID-19 and familiarity with the debunking campaign. | Exposure to seven infographics from the German Ministry of Health. On each infographic, a piece of misinformation was introduced and refuted with a short debunking message. Control group read description of procedure of getting vaccination. | ***Psychological:*** Misinformation belief  ***Behavioural:*** Intentions to get vaccinated against COVID-19. | - A debunking effect on misinformation belief was only found in participants with a medium a priori misinformation belief and did not extend to these participants' vaccination intentions. - Among participants with a high a priori misinformation belief, there was a small unintended backfiring effect on vaccination intentions |
| Hing (2022) *Malaysia* | Online RCT. 14-arm randomised controlled experiment with a parallel design. | Eligible participants were adult Malaysians aged 18 years and above who could understand either the English or Malay language and had not received any dose of the COVID-19 vaccine (n=5784). | Exposure to either one or two messages from a selection of eight different types of messages. The source of the information displayed is stated below the message’s content to provide information credibility. | ***Behavioural:*** Intent to accept the COVID-19 vaccine, intent to recommend the vaccine to healthy adults, to the elderly, and to people with pre-existing health conditions. | - None of the messages improved vaccination intentions. - Intention share information with people with pre-existing conditions improved in descriptive norms and positive attribute frames. |
| Igartua (2023)  *Spain* | Online RCT experiment with a 2 x 2 between-subject factorial design. | 278 adults aged between 18 and 28, who were residing in Spain and had not had a past or current COVID-19 infection. | Narrative messages in the form of a Twitter thread describing a COVID-19 infection (with mild or severe symptoms) that affected either the protagonist of the message (a 23-year-old young person) or their father. | ***Psychological: I***dentification with the protagonist, Narrative transportation , Cognitive elaboration, Reactance (hypothesised mediating variables) Perceived personal risk of COVID-19 infection, perceived severity of COVID-19.  ***Behavioural:*** Protective behavioural intent against COVID-19. | - Severe framing increased perceived personal risk (through indirect effects mediated by cognitive elaboration) and greater intention to adopt preventive behaviors (mediated by both cognitive elaboration and reduced reactance) - Identification with the protagonist and narrative transportation did not constitute relevant mediating mechanisms. |
| Koban (2023) *USA* | RCT with 2 groups - intervention and control. | 371 unvaccinated adults living in the United States, who were Facebook users. | Facebook discussion group with daily posts from group admins covering a variety of topics (e.g. COVID-19 risks, vaccination benefits, and vaccine safety). | ***Psychological:*** Vaccine confidence beliefs, trust in health organisations as mediator.  ***Behavioural***: Intentions to vaccinate, intentions to encourage others to vaccinate. | - Significant interaction between the intervention and trust in public health institutions (PHIs) for improving intentions to vaccinate, intentions to encourage others to vaccinate, and vaccine confidence beliefs. - Among non-conservatives, participants in the intervention had higher posttest intentions to vaccinate. |
| Kopelowicz (2023) *USA* | RCT with two groups – intervention and control | 100 participants who: self-identified as Latinx, spoke Spanish fluently, were between the ages 18 and 74, had a clinical diagnosis of a schizophrenia-spectrum disorder made by a licensed clinician, and had the ability and cognitive capacity to provide fully informed consent. | A 10 min animated, Spanish-language audio-visual novela designed to improve knowledge, attitudes, and behaviours regarding COVID-19. | ***Psychological:*** Knowledge of COVID-19, attitudes including perceived likelihood of infection, perceived effectiveness of safety behaviours.  ***Behavioural:*** Engaging in protective behaviours including mask wearing, frequent hand washing, social distancing, avoid contact with people at high risk and discussing safe practices with a relative. Likelihood of getting vaccinated. | - One month after watching the audio-visual novela, subjects endorsed a greater likelihood of seeking a COVID-19 vaccine than control subjects. - No other significant differences were observed between the two conditions. |
| Legate (2022) *89 countries* | Online RCT, 3 groups. | 25,718 participants from 89 countries. Nil information provided about other inclusion/exclusion criteria or recruitment process. | Exposure to an autonomy supportive message, a pressuring message, or no message. | ***Psychological:*** Autonomous and controlled motivation; feelings of defiance.  ***Behavioural:*** Short- and long-term behavioural intentions – intention to socially distance in the next week and intention to continue socially distancing for the majority of the next six months. | - Those in the no-message condition showed lower controlled motivation than those in the controlling message condition. - The autonomy-supportive message led to lower feelings of defiance than the controlling message, however no difference between the no-message and the controlling message conditions. - Autonomy-supportive condition and Controlling condition did not impact intention to social distance. |
| Li (2023) *China* | Online RCT. 2 intervention and 1 control group. | 981 Chinese citizens aged 18 years and above who had not been vaccinated against COVID-19. | Gain and loss framed messages about getting vaccinated for COVID-19. | ***Behavioural:*** Willingness to vaccinate themselves, their children, and elderly members. | - The gain- and loss-framed messages increased people's willingness to vaccinate themselves, their children, and the elderly. - Compared to the gain-framed messages, the loss-framed messages had a greater impact on enhancing people's willingness to self-vaccinate, but not on vaccinating their children and the elderly. |
| Li (2024)  *Australia* | Online RCT experiment with 3 arms – intervention, usual care and control. | 226 adults who were able to read and understand English, and resided in Australia, recruited through a market research platform. | Intervention arm received diagnostic accuracy information from a community-based study of self-testing provided in a way that was health literacy sensitive. Usual care arm had manufacturer supplied diagnostic information provided. | ***Psychological:*** Understanding of diagnostic accuracy information (RAT sensitivity and specificity)  ***Behavioural***: Intention to self-isolate in 5 hypothetical scenarios. | - More participants in the intervention group correctly interpreted the meaning of the diagnostic accuracy information - The proportion who would self-isolate was similar across scenarios 1 to 3 (likely true positives). - The proportion was higher in the intervention group than in the control for scenarios 4 and 5 (likely false negatives). - However these differences were not statistically significant. |
| Lungu (2023)  *Norway* | Online randomised experiment with 3x2x2 between-subjects design. | 1192 members of the Norwegian Air Ambulance Foundation (NAAF) aged 18 to 90 years. | Participants were show one of twelve short health communication videos related to pandemics. Three factors were included in the creation of the videos: the topic (exponential growth, handwashing, and burden of pandemics on the health care system), the source (expert and nonexpert), and a call to action (present or absent). | ***Psychological:*** comprehension, trust  ***Behaviour***: Past and future intentions, and a proxy for behaviour | - The 3 pandemic-related topics did not affect trust, comprehension, intentions, or behaviour. - Trust was positively influenced by an expert source, whereas a nonexpert source instead had a positive effect on the proxy for behaviour compared with the expert source. - The inclusion of a call to action had a positive effect on both trust and comprehension. |
| Massey (2022) *USA* | Online RCT. 3 intervention groups and control group. | 1004 U.S. individuals residing the U.S., aged 18 years old or older, having smoked 100 cigarettes in their lifetime and currently smoking cigarettes every day or some days, and having sufficient English language ability to participate in the study. | Messages in the COVID-19 risk condition focused on disease progression (lung failure, heart damage, and death). Messages in the smoking risk condition described negative health impacts of smoking (lung and heart disease, cancer, and death). The combined risk condition described how smoking makes COVID-19 worse. The control condition showed non-risk messages (e.g., facts about whales). | ***Psychological:*** COVID-19 perceptions (i.e., perceived severity or susceptibility; self-efficacy or response efficacy); COVID-19 conspiracy beliefs  ***Behavioural:*** COVID-protective intentions (wear mask, wash hands, social distance) | - Highlighting salience of consequences (i.e., underscoring how smoking makes COVID worse) in conjunction with information about health consequences of smoking and COVID increased mask wearing intentions compared with no risk message. - Conditions which provided information about smoking risk or COVID risk alone did not increase mask wearing intentions compared with no risk message. |
| Merle (2024) *USA* | Online randomised experiment with a 3 group between-subjects design. | 350 adults aged 18 years or above, recruited through Amazon’s MTurk. 83.80% reported having a college education and above. | Participants were randomly assigned to view a text, video, or infographic explaining how to wash their hands adequately based on CDC guidelines. | ***Behaviour:*** Intention to practice handwashing guidelines. | - The intention to practice handwashing guidelines did not differ between conditions. - Perceived severity of the risk, perceived benefits of proper handwashing, high self-efficacy towards the behaviour, and cues to action to engage in the behaviour were all significantly related to a greater intention to engage in the suggested handwashing behaviour guidelines. |
| Motta (2021) *USA* | Online RCT. 3 x 2 x 2 design. | 7064 adults living in the U.S.; demographically representative sample. | Pro-vaccine communication materials emphasizing either 1) personal health risks, 2) economic costs, or 3) collective public health consequences of not vaccinating. Also randomly varied the message source (ordinary people vs. medical experts) and availability of information designed the “pre-bunk” potential misinformation about expedited clinical trial safety. | ***Behavioural:*** Intention to vaccinate against COVID-19. | - Messages emphasizing the personal health risks and collective health consequences of not vaccinating significantly increase Americans’ intentions to vaccinate. - These effects are similar in magnitude irrespective of message source, and the inclusion of pre-bunking information. - Economic cost frames have no discernible effect on vaccine intention. - These effects are no different for Democrats, Republicans, and Independents alike. |
| Okuhara (2020) *Japan* | Online RCT. 5 intervention groups and control group. | Men and women aged 18–69 years ( n = 1980). Excluded were criteria were individuals who answered that they cannot go out because of illness or disability; that they have been diagnosed with a mental illness; or/and that they or their family members have been infected with COVID-19. | Five intervention messages: from a governor, a public health expert, a physician, a patient, and a resident of an outbreak area. The content of each message encouraged readers to stay at home and included threat and coping messages. | ***Psychological:*** Perceived severity, response efficiency and self-efficacy.  ***Behavioural:*** Intentions to stay at home. | - Compared with other messages, the message from a physician significantly increased participants' intention to stay at home in areas with high numbers of people infected3 |
| Okuhara (2022) *Japan* | Online RCT. 2 intervention groups and control group. | Men and women with children under the age of 11 (n = 969), who were identified as vaccine hesitant. 77% of the sample was female. | Intervention messages derived from news articles, using a patient’s narrative that targeted the fundamental motive of kin care and that targeted the fundamental motive of disease avoidance. | ***Psychological:*** attitude toward vaccination  ***Behavioural:*** Intention to get vaccinated. | - An intervention message targeting the fundamental motive of kin care and disease avoidance significantly increased intention of vaccination versus a control message. |
| Reddinger (2022) *USA* | Online experimental study. Unclear how many groups. | Americans who had not received a COVID vaccination and who passed an attention check (n=2,621). | Messages that presented the health risks of COVID-19 to oneself and others; they also received messages about the benefits of a COVID-19 vaccine and an endorsement by a celebrity. Messages were randomly tailored to each participant’s identities—Black, Latinx, conservative, religious, or being a parent. | ***Psychological:*** COVID-19 vaccine hesitancy | - No support for the hypothesis that customized messages or endorsers reduce vaccine hesitancy among our segments. - Unregistered post hoc analysis finds evidence that a vaccine endorsement from Dr. Fauci reduces stated intent to vaccinate among conservatives. |
| Ryan (2023) *USA* | RCT in community setting, 2 groups (intervention and control). | 89 residents of La Habra, California, participating in the senior food assistance programs. | All participants received a COVID-19 flyer with clear information on staying safe and resource numbers. Intervention group also received calls over the four months from a health coach for ongoing support. | ***Psychological:*** COVID-19 knowledge.  ***Behavioural:*** COVID-19 mitigation behaviour, Vaccination rates collected at the end of the study. | - No change was observed in COVID-19 knowledge at the end of the study. - COVID-19 mitigation behaviour (composite score) was statistically higher for those adults assigned to intervention versus those assigned to the control. - Significantly higher rate of COVID-19 vaccination in the intervention group compared to and control group. |
| Saling (2024) *Global* | Online experiment with two groups (intervention and control). | 213 participants with a range of vaccine attitudes, recruited from a wide range of apolitical, online groups whose function is to provide information about COVID-19 and its vaccines. | Intervention group was exposed to an accuracy-sensitisation prime. | ***Behavioural:*** Intention to share information online. | - Participants sensitised to accuracy were significantly more likely to share pro vaccine information - However, accuracy-sensitisation had no effect on anti-vaccine information sharing. - The likelihood of sharing anti-vaccine information was positively predicted by the percentage of one's anti-vaccine friends, the size of one's social network, and conservative political orientation. - Conversely, the likelihood of sharing pro-vaccine information was positively predicted by the percentage of one's pro-vaccine friends, and liberal political orientation. |
| Sayfi (2024) Global | Online allocation-concealed, blinded, controlled superiority trial. | 488 individuals from around the world who self-reported to be at least 21 years old, had access to the internet, and were able to read and understand English. | Plain Language Recommendation (PLR) of two COVID -19 recommendations - one from the World Health Organization (WHO) and one from Centers for Disease Control and Prevention (CDC). Control was the Standard Language Version (SLV). | ***Implementation:*** accessibility, usability, satisfaction, preference. ***Psychological:*** Understanding of recommendations. ***Behavioural:*** Intention to follow the recommendation, and intention to share with others. | - Participants in the PLR group had higher understanding scores related to the WHO recommendation, but the difference was not significant for the CDC recommendation - Participants found the PLRs more accessible, more usable and more satisfying. - Participants in the PLR group were more likely to follow the recommendation if they had not already followed and share it with other people. - No significant difference in participants’ preference between different formats. |
| Smith (2024)  *South Africa* | RCT in community setting, 2 groups (intervention and control), cluster randomization by household. | 81 newly diagnosed COVID-19 cases who were over 18 years, and 245 of their household contacts who were over 12 years of age, recruited from two densely populated, low socioeconomic Cape Town community sub-districts. | Infection mitigation intervention delivered by lay community health workers (CHWs) - mitigation measures included one initial household assessment conducted by a CHW in which face masks, sanitiser, bleach and written information on managing and preventing spread were provided, followed by regular telephonic follow-up from CHWs. | ***Health***: SARS-CoV-2 antibodies in HHC participants. | - Participants had significant exposure to SARS-CoV-2 infections prior to the study. In this setting, household transmission mitigation was ineffective. |
| Thaker (2021) *New Zealand* | Online experimental study. 3 intervention groups. | 1083 adults recruited with quotas to be representative of the NZ population. Sample demographics closely matched with the Census estimates on gender, age, and ethnicity but had a higher proportion of educated and fewer Pasifika respondents. | Exposure to vaccine misinformation (a social media post that the vaccine will interfere with genetic material), hesitancy (a newspaper article detailing a mother’s concern to vaccinate son against COVID-19) and factual information (a social media post from a government Facebook page, with information about importance of vaccine for protecting community and economy from COVID-19 outbreaks). | ***Psychological:*** Believability of Information ***Behavioural***: Change in vaccination intention, intention to fact check the information presented, and intention to share the image with friends/followers. | - Exposure to outright COVID-19 vaccine misinformation as well as exposure to vaccine hesitancy induced a decline in COVID-19 vaccination intentions and share information with friends compared to control or factual information. - Exposure to misinformation and hesitancy reduced believability in presented information, with hesitancy condition more likely to believe than misinformation condition. - Respondents were more likely to believe in vaccine hesitancy information and share such information with family and followers compared to misinformation. |
| Thunstrom (2021) *USA* | Between-subjects experimental design with eight information treatments (2 × 2 × 2). | 3,133 respondents representative of the US general population in gender, age, income, education, race, and residential region." | The experiment varied information on (1) the probability of the average American catching the coronavirus, (2) the IFR, i.e., the probability of the average American dying if infected, and (3) the source of information for the probability of catching COVID-19 (CDC only/CDC jointly with the White House)." | ***Psychological:*** Perceived susceptibility, perceived severity  ***Behavioural:*** Intention to vaccinate. | - Vaccination intention was lower in the low-risk probability condition and inconsistent information from official sources condition. - Perceived risk was lower in the low-risk probability condition |
| Torres (2021) *USA* | Online RCT. 2x2 design. | 18223 participants aged 18 years or older, self-identifying as White or Black, and without a college degree, | Video messages on COVID-19 (including common symptoms, case numbers and CDC social distancing guidelines) delivered either by a Black or a White study physician, and an American Medical Association written statement on structural inequity. | ***Psychological***: COVID-19 knowledge  ***Behavioural***: Information seeking behaviour, self-reported protective behaviours, willingness to pay for a mask. | - Compared with the control group, the intervention group had smaller gaps in COVID-19 knowledge and greater demand for COVID-19 information, and willingness to pay for a mask. - Self-reported safety behaviour improved, although the difference was not statistically significant - The intervention was impactful for both Black and White participants. - It was more impactful for White participants vs Black participants on knowledge but equally impactful for all the other measures |
| van Empelen (2022) *Netherlands* | Online RCT. 3 experiments each with 1 intervention and 1 control group. | Per experiment 424-593 adults aged 18 years or older who lived in the Netherlands and mastered the Dutch language. | Volitional Help Sheet: with “if-then” statements, about potential difficult situations to comply with the COVID-19 precautionary measures and corresponding possible solutions. Behavioural Journalism: Short films of individuals sharing the impact that COVID-19 has on their lives and how they dealt with it; role-modeling precautionary measures and explaining why they believe it is important to comply. Empathy Induction: Short film depicting an at-risk person explaining her dependency on others following precautionary measures to be protected. Participants who indicated intention to protect others were offered a gift. | ***Psychological:*** Self-efficacy and intention to comply with COVID-19 precautionary measures, perceived susceptibility to get infected with COVID-19, perceived severity to get infected with COVID-19, perceived susceptibility of others to get infected with COVID-19, response efficacy to reduce the risk of infecting oneself and others.   ***Behavioural:*** Intention to comply with COVID-19 precautionary measures, subjective behavioural compliance with COVID-19 measures. | - Two out of the three different strategies did result in favourable changes with regard to the compliance-related measures. - The VHS contributed to changes in perceived susceptibility of others and individual behavioural compliance measures. - People exposed to the VHS were more likely to receive less visitors and avoid crowds. - EI increased the perceived vulnerability of others. - Video-based role model stories, based on BJ did not result in any changes. |
| Vandormael (2021) *USA, Mexico, UK, Germany, Spain* | Online RCT with 3 groups – intervention, attention placebo control, do-nothing. | 14,482 adults aged 18 to 59 years who were residents of one of the five countries, and had proficiency in English, German, or Spanish | CoVideo is short animated video with sound effects. It explains how COVID is transmitted and recommends best practices to prevent onward transmission (staying at home, not congregating in public spaces, and sanitizing hands/surfaces). It includes a subplot on the stockpiling of essential goods, and the impact thereof on health care services and resources (e.g., doctors being unable to access protective equipment). | ***Psychological:*** COVID-19 knowledge ***Behavioural***: Behavioural intent to go out with friends, to wash dishes, wash hands frequently, clean kitchen counters, and stockpile household supplies. | - Knowledge in the CoVideo arm was significantly higher than in the do-nothing arm. - High baseline levels of behavioural intent to perform many of the preventive behaviours featured in the video intervention were observed. - We were only able to detect a statistically significant impact of the CoVideo on one of the five preventive behaviours (intention to wash dishes), when compared to the passive control. |
| Vivion (2022) *Canada* | Online RCT. 3x2x2 design. | 2500 participants aged 50 years and older, understanding French or English, with internet access who had not yet received any COVID-19 vaccination. | Inoculation intervention - warning message of an impending threat/attack on one’s prior belief/attitude and 2 different disinformation messages (on mRNA and quick approval of vaccines). | ***Psychological:*** Changes in attitudes toward COVID-19 vaccination.  ***Behavioural:*** Intention to get vaccinated against COVID-19. | - No meaningful results in regards to attitudes. - Group comparisons between those who received only disinformation and those who received the inoculation message show that prebunking messages may safeguard intention to get vaccinated and have a protective effect against disinformation. |
| Yildirim (2020) *USA* | Between-subjects experimental design with 3 groups. | 2617 US residents aged 18 years or older. | “Equal pandemic” framing, which emphasized how the outbreak had been affecting everyone regardless of their background; the “elderly and medical conditions inequality” framing, which emphasized that the pandemic had been especially hard on the elderly and those with medical conditions; and the “class inequality” framing, which specifically emphasized that the pandemic had been especially hard on the poor and low-income communities. | ***Psychological:*** opinions regarding whether coronavirus is a serious threat or not are concerned; opinions as to whether the priority should be saving lives or saving the economy. | - The elderly and medical conditions inequality group reported significantly lower levels of threat perception compared to the equal pandemic group. - The elderly and medical conditions inequality condition also reported significantly more support towards saving the economy over saving lives compared to the equal pandemic condition. |
| Yu (2022) *China* | RCT with 2 groups - intervention and control. | 296 Chinese residents aged 18 years or older who were vaccinated against COVID-19 and were able to read or understand the study. | An educational leaflet comprising of two sections: 1) COVID-19 knowledge included the clinical manifestations, transmission routes, and prevention and control strategies of COVID-19 and 2) Vaccine knowledge included China's COVID-19 vaccination policy, indications and contraindications, adverse reactions, effects, and postvaccination prevention and control measures. | ***Psychological***: Knowledge of COVID-19 and vaccination. | - Reading leaflets during the observation period after vaccination effectively improved their knowledge. |
| Yuan (2022) *USA* | Online RCT, 3 intervention groups and control group. | 702 participants residing in the US who had not yet received a COVID-19 vaccination. | Four short videos encouraging viewers to get vaccinated with the COVID-19 vaccine. The individual-centred message focused on the benefit of the vaccine on protecting themselves. The community-centred message focused on the vaccine’s ability to protect the community in which the viewers live. The country-centred message illustrates the country-level benefit of the COVID-19 vaccine. Control video had no specific message. | ***Psychological:*** Support for vaccine mandate.  ***Behavioural:*** Willingness to vaccinate. | - Respondents were more likely to get vaccinated and support vaccine mandates after viewing an individual-centred message, less with a community-centred message. - Individuals who value individualism were more likely to respond positively to individual-centred messages, but those who believe more in communitarianism value were less likely. |
| Yuen (2023) *Hong Kong* | Online RCT with 3 groups. | 1072 adults in the Hong Kong general population | Vignettes with a neutral government announcement only; vignettes with a government announcement and a health expert's quote supporting the government's decision; and vignettes with a government announcement and a health expert's quote disagreeing with the government's decision. | ***Psychological:*** Support for COVID-19 measures (banning public assembly, contact-tracing mobile applications and restriction testing), trust in health experts. | - Positive health experts' communication increased the support for banning public assembly; no effects were found for the support for contact-tracing mobile applications and restriction testing. - Participants who only viewed health experts disagreeing with the government had higher trust in health experts relative to participants who viewed health experts agreeing with the government at least once. - The results render doubtful the strategy that health experts can be involved for garnering support for unpopular health measures without jeopardizing public trust in them. |
| Zhang (2024)  *China* | RCT in school setting, 2 groups (intervention and control), cluster randomization by class. | 2526 third-grade classes from 11 pilot schools in Longgang district of Shenzhen, China | Two free 40 minute sessions of health education during the study – including a lecture on transmission and prevention of different infectious diseases, followed by a 5-minute science video, with quizzes and prizes to incentivise learning. | ***Psychological:*** Knowledge of infectious disease (including COVID-19) symptoms and transmission.  ***Behavioural:*** COVID-19 vaccination rates | - After the intervention, students in the intervention group had significantly greater knowledge and also greater improvement in knowledge scores than the control group. - Differences in vaccination rates were not significant between the two groups. |
| Zhao (2024)  *China* | Online randomised experiment, 2x2 between-subjects. | 371 adults recruited from an online survey platform in China. | Focusing on the discussion of whether the asymptomatic cases detected during the COVID-19 pandemic would further lead to an uncontrolled pandemic, news articles were manipulated in terms of whether the infectiousness of asymptomatic cases and the means to control the transmission are presented in terms of their certainty or uncertainty. | ***Psychological:*** Vulnerability to False Information, Perceived Severity, Perceived  Susceptibility  ***Behavioural:*** Willingness to Adopt Preventive Behaviours, Information Avoidance, Information Seeking | - Individuals were more susceptible to believing false COVID-19-related information when a certain threat and uncertain solution were framed in the news article. - Perceptions of crisis severity increased when exposed to news information containing uncertain solutions. - Information seeking was positively associated with protective behavioural intention, whereas information avoidance was negatively associated with protective behavioural intention. |
